# Optimal Pool Size for COVID-19 Group Testing

**DOI:** 10.1101/2020.04.26.20076265

**Authors:** Jeffrey Y. Chen, Andrew S. C. Chen

## Abstract

This paper presents an analytical formulation for determining optimal pool size in the initial pooling stage and the subsequent retests for COVID-19. A generalized constant compaction approach confirms the efficiency of “halving” targeted population between retest stages. An analytical gain formula is derived to aid future test designs. It is observed that optimal gain relies on the proper choice of the initial pool size. This optimal compaction scheme outperforms the conventional algorithms in most cases and may provide a mathematically-native road map for us to operate beyond the standard super-even-number-based (64, 32, 16, 8…, 1) group testing algorithms.

## 1. Introduction

Group (or pooled) testing for a disease (or other attribute) involves testing “pools” of individual samples. If a pool tests negative, then all samples therein are assumed negative; if it tests positive, then further pooled or individual tests ensue. The approach was first suggested by Dorfman in 1943 [1] and has been greatly developed over the years [2–4]. The game of improving group testing efficiency is largely driven by the prevalence^1*^ rate. The strong appeal of pooled testing is that it can significantly reduce the number of tests and associated costs when the prevalence for a disease is small. The 2020 COVID-19 pandemic calls for timely extensive testing for a large populace. The lack of available tests aggravates the situation around the world, and hence this study. At the time of writing, Germany [5] and Israel [6] have reported group testing efforts. This study aims to develop a strategy for choosing the optimal pool size and compaction factor for each stage to further improve the testing efficiency. Instead of starting with a super-even number (4, 8, 16, 32, etc.) as the pool size, the approach developed in this paper computes the optimal pool size based on the prevalence information dynamically for every stage. The pooled (and hence compacted) prevalence is used in determining the pool size for the next stage. The strategies for the conventional approach and the Optimal Pool Size approach are illustrated in Figure 1.

**Figure 1.**
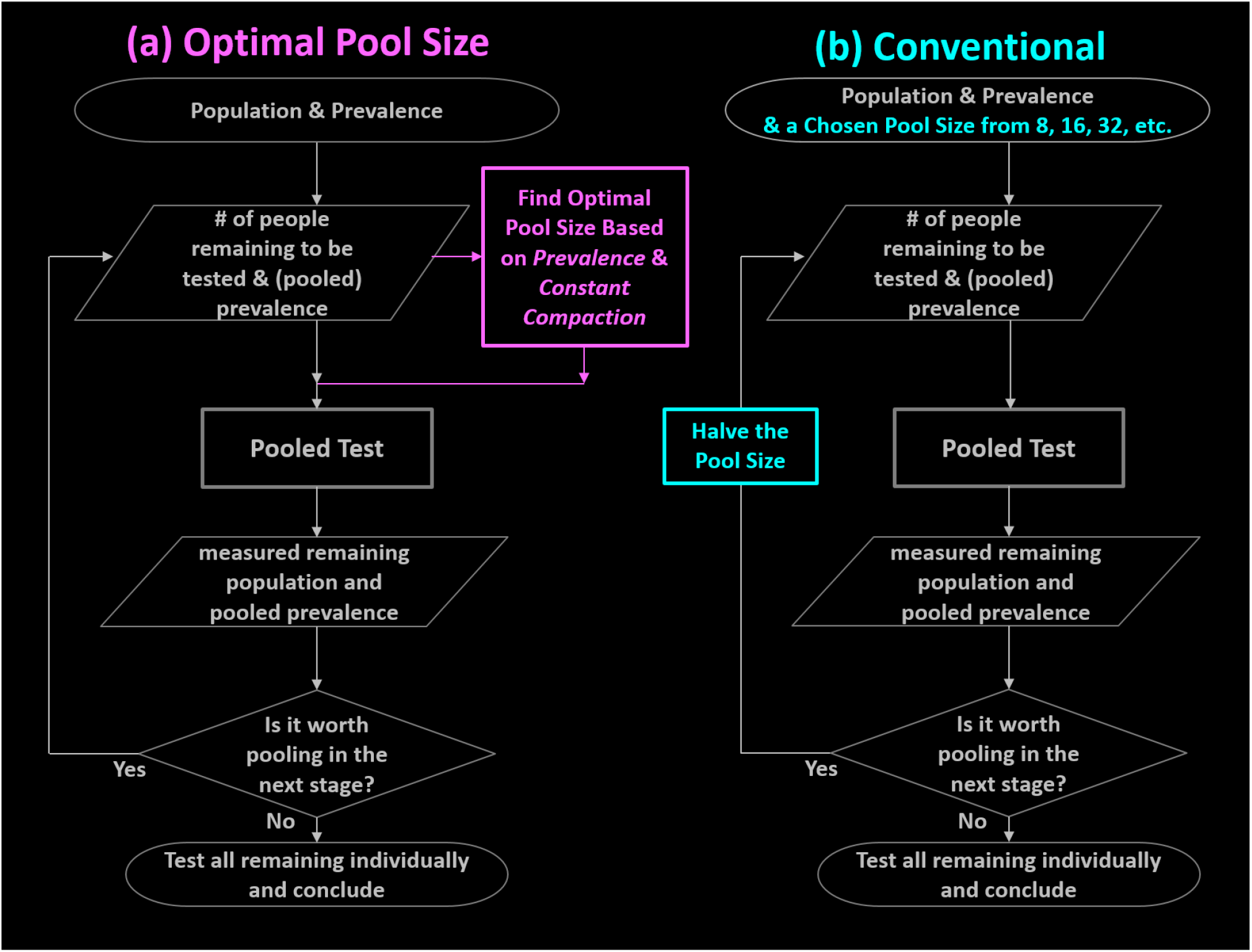
Comparison of the flowcharts for (a) the Optimal Pool Size approach, and (b) the conventional approach. The difference is highlighted in color: The Optimal Pool Size approach (magenta) dynamically computes the optimal pool size based on the knowledge of prevalence. The pooled (and hence compacted) prevalence is used in determining the pool size for the next stage. In contrast, the conventional approach (cyan) halves the pool size for each successive stage.

Since the algorithm under study searches and then uses an optimal group size at every pooling stage, it outperforms conventional approaches consistently. Figure 2 serves as a visual synopsis of this observation. In Figure 2, the gain in testing efficiency of this approach is used as the benchmark, set equal to 1, for a prevalence ranging from 0.1% to 30%. It is clearly seen that the *Optimal Pool Size* approach provides better gain values. Cases with initial pool size of 8, 16, 32, and 64 take turns approaching the efficiency of the Optimized Pool Size approach. The colored diamonds indicate locations of prevalence values that lead to super-even number optimized pool sizes, i.e., where conventional super-even-number based systems approach and become the same as the Optimized Pool Size design. For example, the green diamond’s prevalence value is 8.66%, the corresponding Optimal Pool Size is 8 because −log(0.5)/0.0866 = 8, according to Eq. 10.

**Figure 2.**
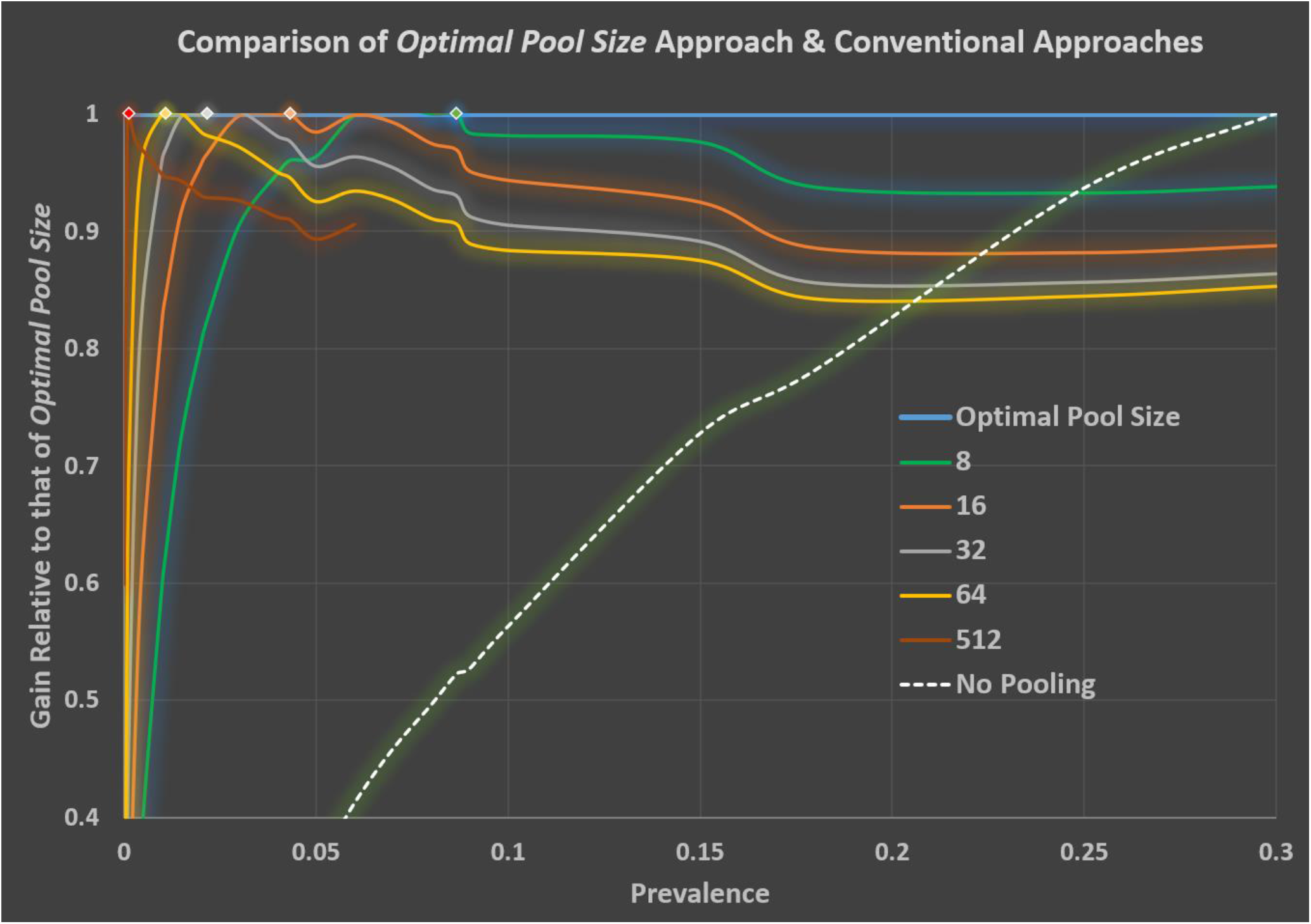
Since an optimal group size is dynamically calculated and applied at each pooling stage, the Optimal Pool Size approach outperforms conventional approaches consistently. The approach’s testing efficiency gain, normalized to one, is used as the benchmark for comparison with conventional approaches. The prevalence values range from 0.1% to 30%. It is clearly seen that the Optimal Pool Size approach provides better gain values than conventional approaches. The colored diamond points illustrate the prevalence values corresponding to the super-even numbers and hence the values of the gain approach that of the optimal Pool Size. For example, the green diamond’s prevalence is 8.66%, the corresponding Optimal Pool Size is 8 because −log(0.5)/0.0866 = 8, according to Eq. 10

A mathematical model of this approach is constructed in the following.

## 2. Formulation

The main parameters we will use in the model include: *p*_*i*_: prevalence at stage *i, b*_*i*_: pool size for stage *i, n*_*i*_: the number of individual samples to be tested in stage *i*, and *N*: total number of people in the target population to be tested. It is obvious that *n*_1_=*N*—the full population is targeted in stage 1. The probability of a particular pool tested negative in stage *i* is 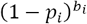. The number of pools tested negative in stage 1 is 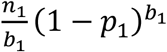. The number of pools tested positive in stage 1 is 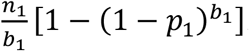. The number of people tested positive after stage *i* is therefore 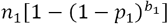. Recall that *n*_1_ *becomes*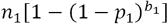 in stage 1. It is productive to define a *Compaction Factor ϵ*_1_for stage 1:

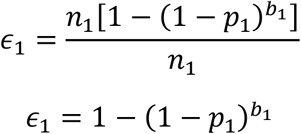

In general, the compaction factor in stage *i* is

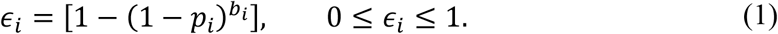

As a result of a compaction *ϵ*_*i*_ in stage *i*

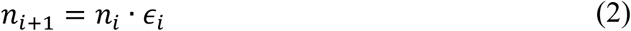

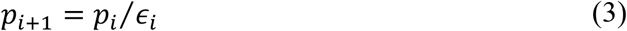

Pooling in a given stage reduces the size of the population to be tested and increases the prevalence in the next stage. The task here is that given *p*_*i*_, one must choose a good combination of pool size *b*_*i*_ and an effective compaction factor *ϵ*_*i*_to drive next stage compaction.

The critical parameters *ϵ*_*i*_ depends on *b*_*i*_ as shown in (1). To see how *b*_*i*_ depends on *ϵ*_*i*_, solve for *b*_*i*_ using (1).

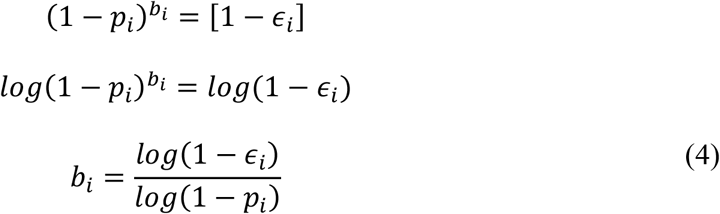

In this paper, all *log* refers to *log*_*e*_. For our region of interest, 0 ≤ *p*_*i*_ ≪ 1, Eq (4) can be approximated with

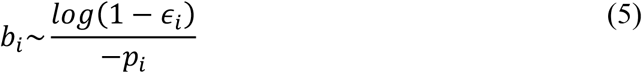

Recall that the goal of group testing is to identify all positive individuals with as few total tests as possible. We need to examine the number of tests required. The number of tests required in stage 1 is 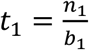. The number of tests required in stage 2 is 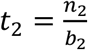, and so on. Denote the total number of stages as *S*. The compaction continues thorough *S*-1 stages until the prevalence rate *p*_*s*_ is so large that it is more effective to just test each sample individually, i.e., set the pool size *b*_*S*_ = 1, and 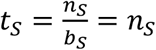. The pool size *b*_i_ is rounded into a natural number in the computation. The total number of tests required is 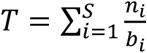.The figure of merit we used to guide the study is the gain *g* defined as

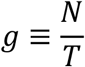

More specifically,

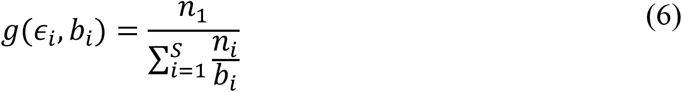

Computer simulations were used to scan through parameter space {*p*_1_, *ϵ*_*i*_, *b*_*i*_}to hunt for effective compacting algorithms. The main results are summarized in Figure 3.

**Figure 3.**
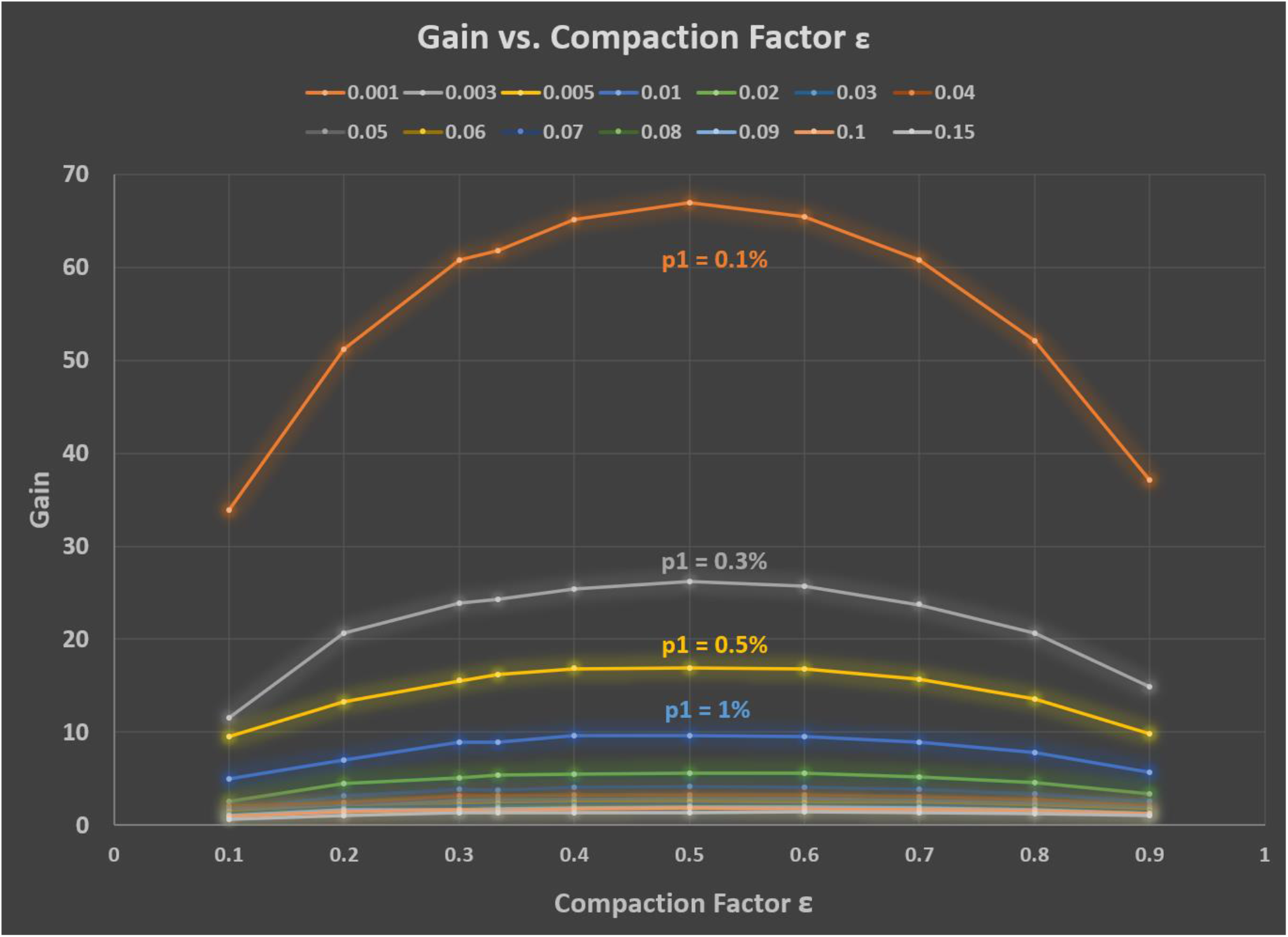
Computer simulation was used to conduct a parameter scan through the “pool size” - “compaction” space {*b*_*i*_, *ϵ*_*i*_} to look for high gains--given various values of prevalence *p*_*1*._ The results revealed a consistent pattern. The best gains are obtained with a constant compaction *ϵ*_*i*_ of 0.5 for all stages, except for the last stage where there is no pooling. The smoothness of the curve, in reality, is modulated by the ceiling function in the gain formula (Eq 16).

Simulation results conclusively indicated that the best strategy for choosing {*ϵ*_*i*_, *b*_*i*_} to maximize the gain *g*is to use a constant compaction of 0.5 in stages 1 through *S*-1.

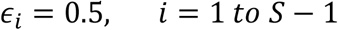

Deploying all *ϵ*_*i*_ with an identical value of 0.5 is a clear indication that this algorithm is structurally a close cousin of the standard 64-32-16-8 “halving” algorithms. The distinctive feature of the approach in this study is the continuous optimization of pool size *b*_*i*_. It determines an optimal pool size at every stage that deviates freely from the super even sequence of “2-4-8-16…” Such an approach regularly outperforms the standard group testing algorithms based on the conventional super-even sequence.

The results of two sample runs are summarized in Table 1: (1) *p* = 1% and (2) *p* = 2%. A population size of 1,000,000 is used. These are the only two input parameters: *p* and *N*.

**Table 1.** Comparing Gain values of Optimal Pool Size Approach with Conventional Approach.

| Prevalence | Optimal Pool Size Approach | Conventional Approach |
| --- | --- | --- |
| 2% | 5.48 | 5.24 |
| 1% | 9.451 | 9.449 |

Table1shows the 2%-prevalence simulation results of a gain of 5.48 using the Optimized Pool Size approach developed here. In contrast, a conventional approach achieved a gain of 5.24. The 1%-prevalence simulation shows a gain of 9.451 using the Optimized Pool Size approach and the conventional approach has a gain of 9.449.

Tables 2 and 3 compare details of the 2%-prevalence simulation runs. Tables 4 and 5 compare details of the 1%-prevalence simulation runs. The pool size progression of 8-4-1 instead of 8-4-2-1 at the late stages is an optimization triggered by a threshold prevalence of p = 0.3034 above which pooling no longer helps. The threshold prevalence will be derived in a later section.

**Table 2.** A gain of 5.48 is achieved for a sample run using the Optimized Pool Size algorithm developed in this study. Input: 2% prevalence, N = 1,000,000. Total Tests Required: 182,619

| stage | population | prevalence | pool size | compaction | population for the next stage | prevalence for the next stage | tests required |
| --- | --- | --- | --- | --- | --- | --- | --- |
| i | $n_i$ | $p_i$ | $b_i$ | $\epsilon$ | $n_{i+1}$ | $p_{i+1}$ | $n_i/b_i$ |
| 1 | 1,000,000 | 2.0% | 34 | 0.497 | 496863 | 4.0% | 29,412 |
| 2 | 496863 | 4.0% | 17 | 0.503 | 249745 | 8.0% | 29,227 |
| 3 | 249745 | 8.0% | 8 | 0.487 | 121662 | 16.4% | 31,218 |
| 4 | 121662 | 16.4% | 4 | 0.512 | 62346 | 32.1% | 30,416 |
| 5 | 62346 | 32.1% | 1 | 1.000 | 0 | NA | 62,346 |

**Table 3.** A gain of 5.24 is achieved for a sample run using the conventional algorithm. Input: 2% prevalence, N = 1,000,000. Total Tests Required: 190,975

| stage | population | prevalence | pool size | compaction | population for the next stage | prevalence for the next stage | tests required |
| --- | --- | --- | --- | --- | --- | --- | --- |
| i | $n_i$ | $p_i$ | $b_i$ | $\epsilon$ | $n_{i+1}$ | $p_{i+1}$ | $n_i/b_i$ |
| 1 | 1000000 | 2.0% | 16 | 0.276 | 276202 | 7.2% | 62,500 |
| 2 | 276202 | 7.2% | 8 | 0.452 | 124821 | 16.0% | 34,525 |
| 3 | 124821 | 16.0% | 4 | 0.503 | 62744 | 31.9% | 31,205 |
| 4 | 62744 | 31.9% | 1 | 1.000 | 0 | NA | 62,744 |

**Table 4.** A gain of 9.451 is achieved for a sample run using the Optimized Pool Size algorithm developed in this study. Input: 1% prevalence, N = 1,000,000. Total Tests Required: 105,809

| stage | population | prevalence | pool size | compaction | population for the next stage | prevalence for the next stage | tests required |
| --- | --- | --- | --- | --- | --- | --- | --- |
| i | $n_i$ | $p_i$ | $b_i$ | $\epsilon$ | $n_{i+1}$ | $p_{i+1}$ | $n_i/b_i$ |
| 1 | 1000000 | 1.0% | 69 | 0.500 | 500163 | 2.0% | 14,493 |
| 2 | 500163 | 2.0% | 34 | 0.497 | 248455 | 4.0% | 14,711 |
| 3 | 248455 | 4.0% | 17 | 0.503 | 124876 | 8.0% | 14,615 |
| 4 | 124876 | 8.0% | 8 | 0.487 | 60832 | 16.4% | 15,609 |
| 5 | 60832 | 16.4% | 4 | 0.512 | 31173 | 32.1% | 15,208 |
| 6 | 31173 | 32.1% | 1 | 1.000 | 0 | NA | 31,173 |

**Table 5.** A gain of 9.449 is achieved for a sample run using the conventional algorithm. Input: 1% prevalence, N = 1,000,000. Total Tests Required: 105,831.

| stage | population | prevalence | pool size | compaction | population for the next stage | prevalence for the next stage | tests required |
| --- | --- | --- | --- | --- | --- | --- | --- |
| i | $n_i$ | $p_i$ | $b_i$ | $\epsilon$ | $n_{i+1}$ | $p_{i+1}$ | $n_i/b_i$ |
| 1 | 1000000 | 1.0% | 64 | 0.474 | 474404 | 2.1% | 15,625 |
| 2 | 474404 | 2.1% | 32 | 0.494 | 234481 | 4.3% | 14,825 |
| 3 | 234481 | 4.3% | 16 | 0.502 | 117731 | 8.5% | 14,655 |
| 4 | 117731 | 8.5% | 8 | 0.508 | 59856 | 16.7% | 14,716 |
| 5 | 59856 | 16.7% | 4 | 0.519 | 31046 | 32.2% | 14,964 |
| 6 | 31046 | 32.2% | 1 | 1.000 | 0 | NA | 31,046 |

The slight deviation of compaction *ε* from 0.5 is mainly caused by the rounding of the computed pool size *b* into the closest integer.

In the following, we will construct an analytical model for the gain values in testing efficiency based on a constant compaction factor *ϵ*.

## 3. Constant Compaction Model

The Constant Compaction Factor will be denoted as *ϵ* whose value is chosen to be 0.5 based on the simulation results. A common compaction *ϵ* in all compaction stages leads to the following.

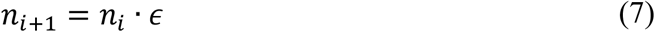

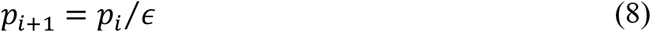

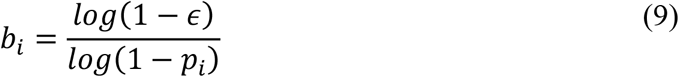

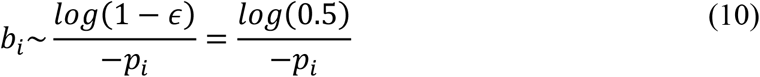

The optimal pool size at every stage depends on the prevalence at that stage. The dependence is shown in Figure 4.

**Figure 4.**
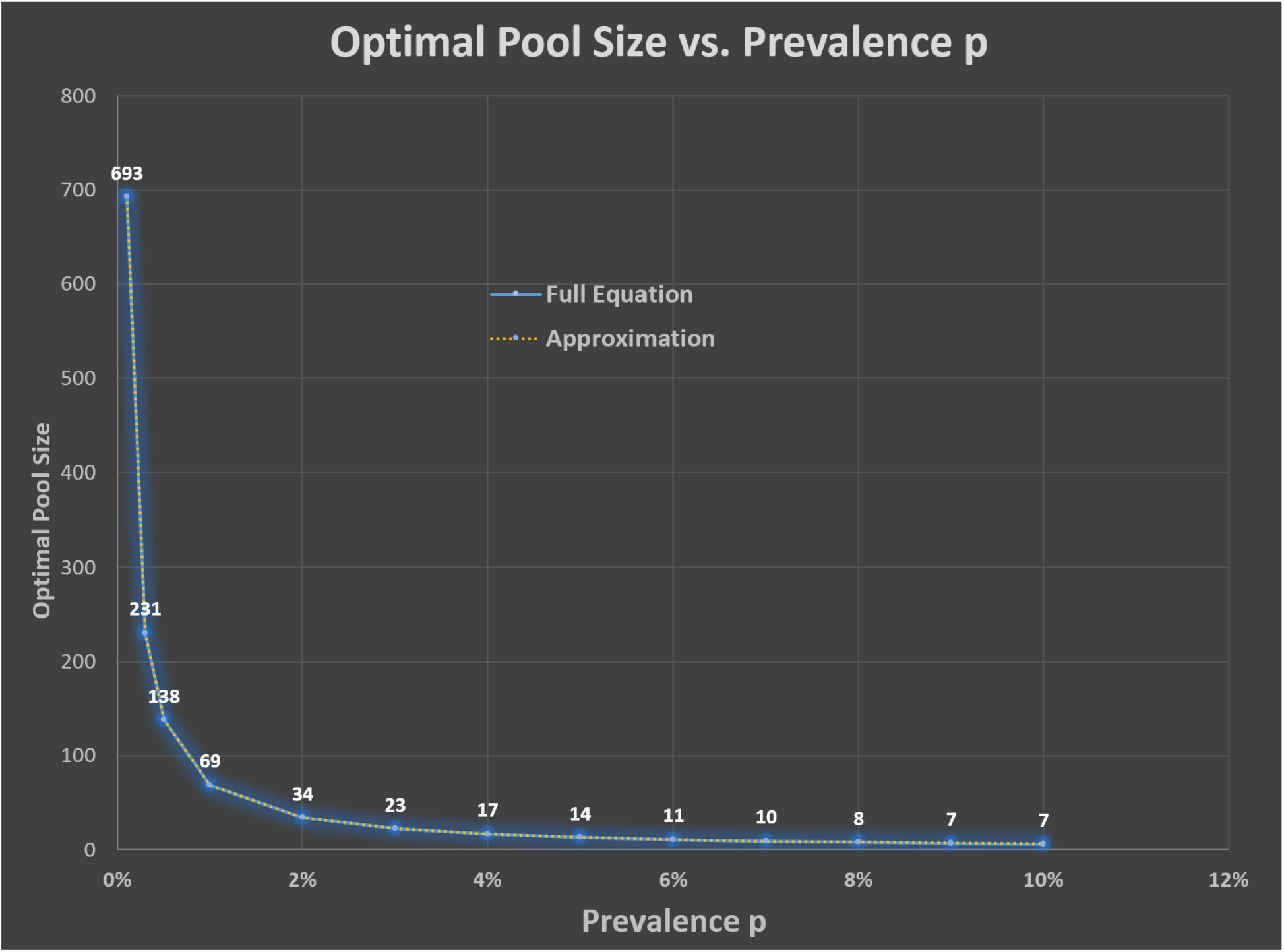
Optimal pool size is plotted against prevalence. The approximated values and the exact values agree well. See Eqs (9) and (10).

It is worth noting that Eq (10) is an almost perfect approximation for Eq (9) in our operating range 0 ≤ *p*_*i*_ ≪ 0.15. [The upper bound of 0.15 is half of the threshold prevalence of 0.3034 which terminates the pooling process. The threshold prevalence will be derived as equation 12.] The error is *O*(*p*_*i*_^2^). It could be used as a simple rule of thumb when one wants to determine the optimal pool size quickly.

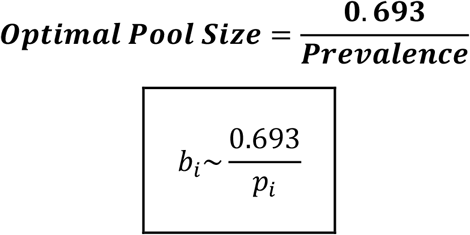

Furthermore, the constant compaction leads to some desirable simplifications: *n*_*i*_=*n*_1·_*ϵ*^*i*−1^ and. Consider the number of tests required in each stage. In stage 1, 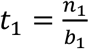, in subsequent retest stages,

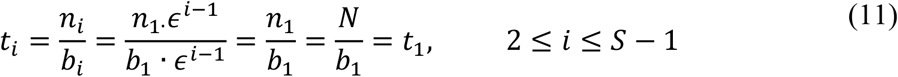

The fact that ***each retest stage requires the same number of tests as the initial stage*** (except for the last non-pooling stage, *S*) simplifies the derivation of the gain in closed form. To calculate the gain, we will need to know the number of pooling stages *S-1*. The very last (“test-every-remaining-sample”) stage is triggered by a *threshold in prevalence*.

At the end of each stage *i* near the end, with a population of *n*_*i*_ remaining to be tested and an expected prevalence of *p*_*i*_, a decision will be made regarding whether to (1) test everyone individually and conclude the test, or (2) keep pooling in the next stage and perhaps wrap up in the stage after that. (1) ***Stop pooling and test everyone***. The required number of tests to conclude the whole test will be *n*_*i*_. (2) ***Keep pooling***. The required number of tests to conclude the test in the last stage will be the *number of pooled tests for the next stage* + *population remained and will be tested individually after that stage*.

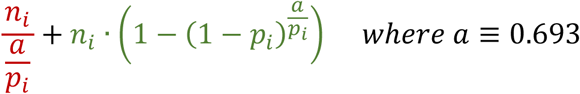

The logic is that we will stop pooling and test everyone, *n*_*i*_, with *n*_*i*_ individual tests if the above expression (for pooling in the next stage) is larger than *n*_*i*_ and therefore the pooling offers no advantage,

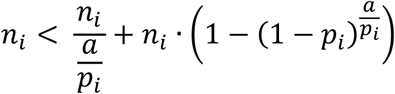

The condition simplifies to

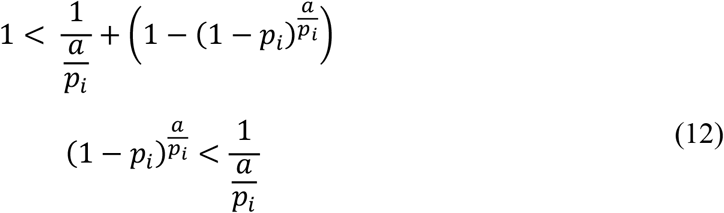

The inequality can be solved graphically for the threshold prevalence *p*_*c*_, yielding *p*_*c*_=0.3034 < *p*_*i*_.

One can also use binomial expansion to solve the inequality analytically with a good approximation.

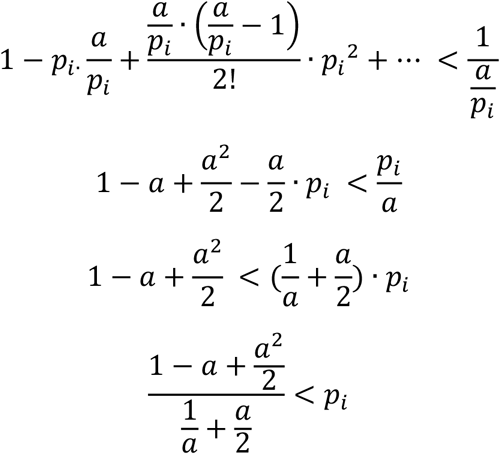

Plugin *a*=0.693,

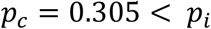

This analytical approximation is very close to the *threshold in prevalence* value 0.3034 obtained from graphical solution of Eq (12). In the following, when the prevalence of the next stage is more than 0.3, or 30%, then we test everyone and conclude the test. Now we are ready to calculate the number of stages required.

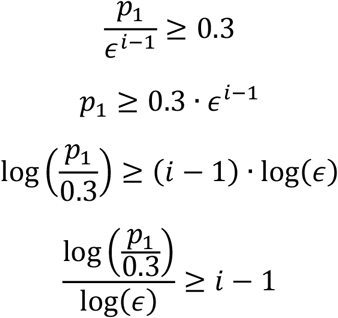

So, the total number of stages with pooling, not counting the very last stage which every sample is tested, is *S*−1

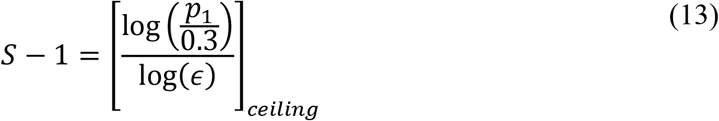

The ceiling function in the equation means rounding up to an integer for obtaining an integer number of stages. As a result, the smoothness of the overall gain formula, in reality, is modulated by the ceiling function. The effect can be seen in Figures 3 and 5. This is also true for the rounding required in Eq (4) for obtaining integer optimum pool size. The effect will be further studied in Section 5.

**Figure 5.**
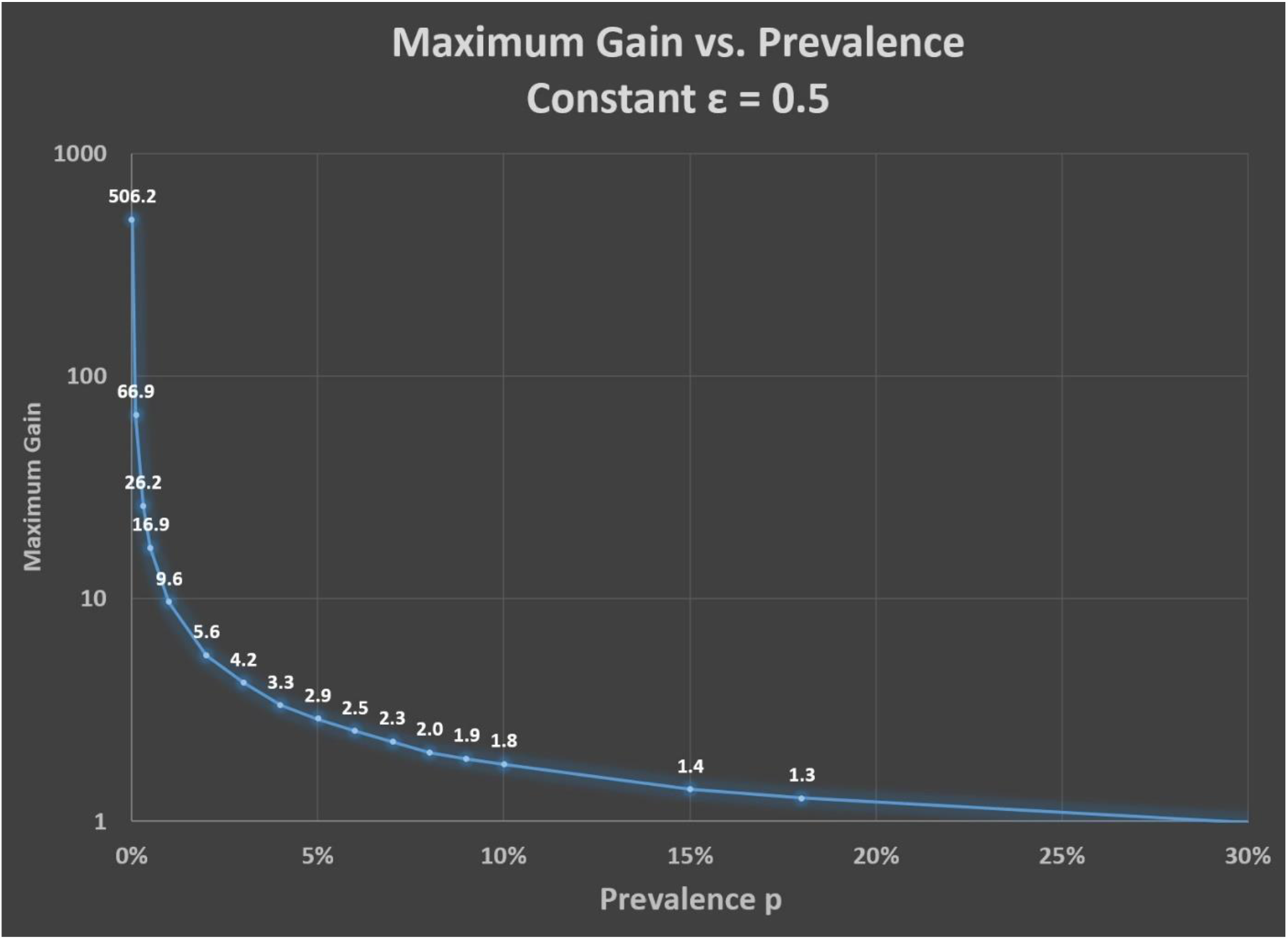
Dependence of maximum gain in group testing efficiency on Prevalence. The smoothness of the curve is perturbed by the ceiling functions in the gain formula. See Eq 16. The high gain regime of operation requires a prevalence lower than 1%. Smart screening and heterogeneous grouping can be used to accomplish high gains. See the example in the next section.

The number of tests in the last stage is

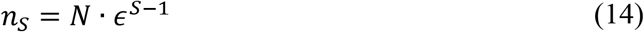

Combining all the required tests for all stages using Eqs (11) and (14), we get the total number of tests

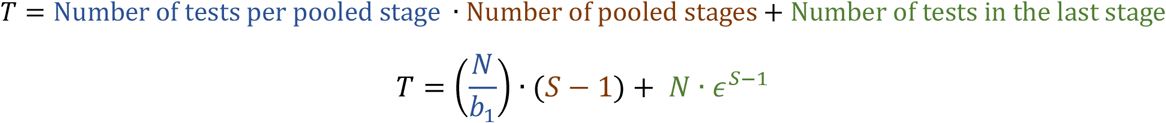

Recalling the gain

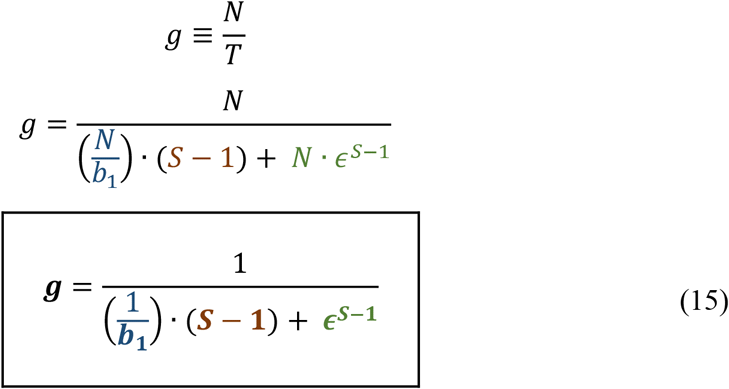

This simple-looking formula for gain elegantly embodies the group-testing mechanism. It captures the dependence of gain ***g*** on the choice of the initial pool size ***b***_1_. Note: 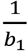, or more precisely, 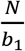, sets the number of tests per stage in the initial and all subsequent pooled stages. Further note: *b*_1_is determined by *p*_1_. The number of pooled-test stages ***S*** − **1** is solely determined by the prevalence *p*_1_ (See Eq 13). Above all, all the elegance and efficiency come from a constant compaction factor ***ϵ***.

A full expression for the gain is

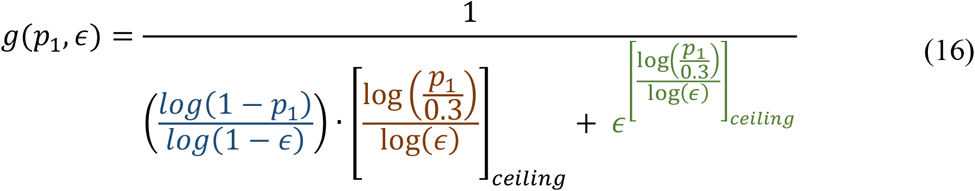

When the *ϵ* in the equation is set to 0.5 in the model, sans the ceiling function, the gain becomes

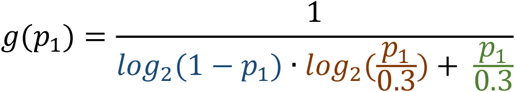

It is clearly seen in the equation that the whole multi-stage group testing optimization through constant compaction is driven by only one critical parameter—the initial expected prevalence *p*_1_. High gains in group testing efficiency can be expected for *p*_1_ ≪ 1. The dependence of gain on prevalence is shown in Figure 5.

## 4. Comparison of *Optimal Pool Size* Approach with Conventional Approach

Based on the model constructed above, a systematic comparison of the available gain for *optimal pool size* approach and the conventional approach was conducted. For the conventional approach, initial pool sizes of 8, 16, 32 and 64 were used with various prevalence values ranging from 0.1% to 30%. The results are summarized in Figure 2 earlier and again in Figure 6 with a semi-log plot.

**Figure 6.**
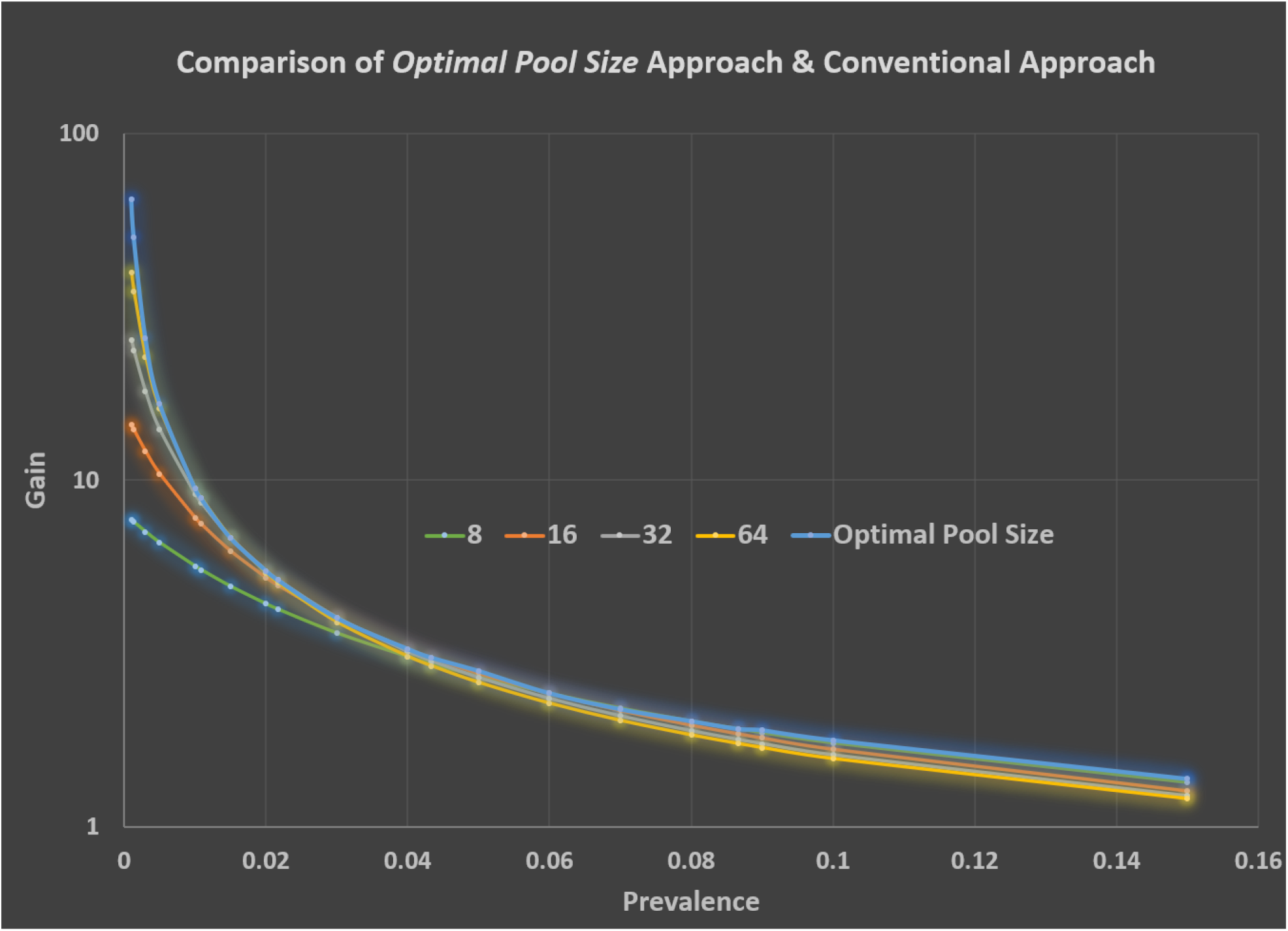
The Optimal Pool Size model developed in this paper consistently outperforms the conventional super-even-number-based approach.

For every prevalence at hand, there is an optimal pool size, as described by *Optimal Pool Size*=(0.693)/*Prevalence*. This important conclusion can be clearly seen when one zooms in to Figure 6. For example, in between a prevalence of 3% and 15%, one can identify the signatures of the super-even numbers16 and 8 using Eq (9) or Eq (10) in Figure 7. Other super-even numbers can be tracked down in other intervals.

**Figure 7.**
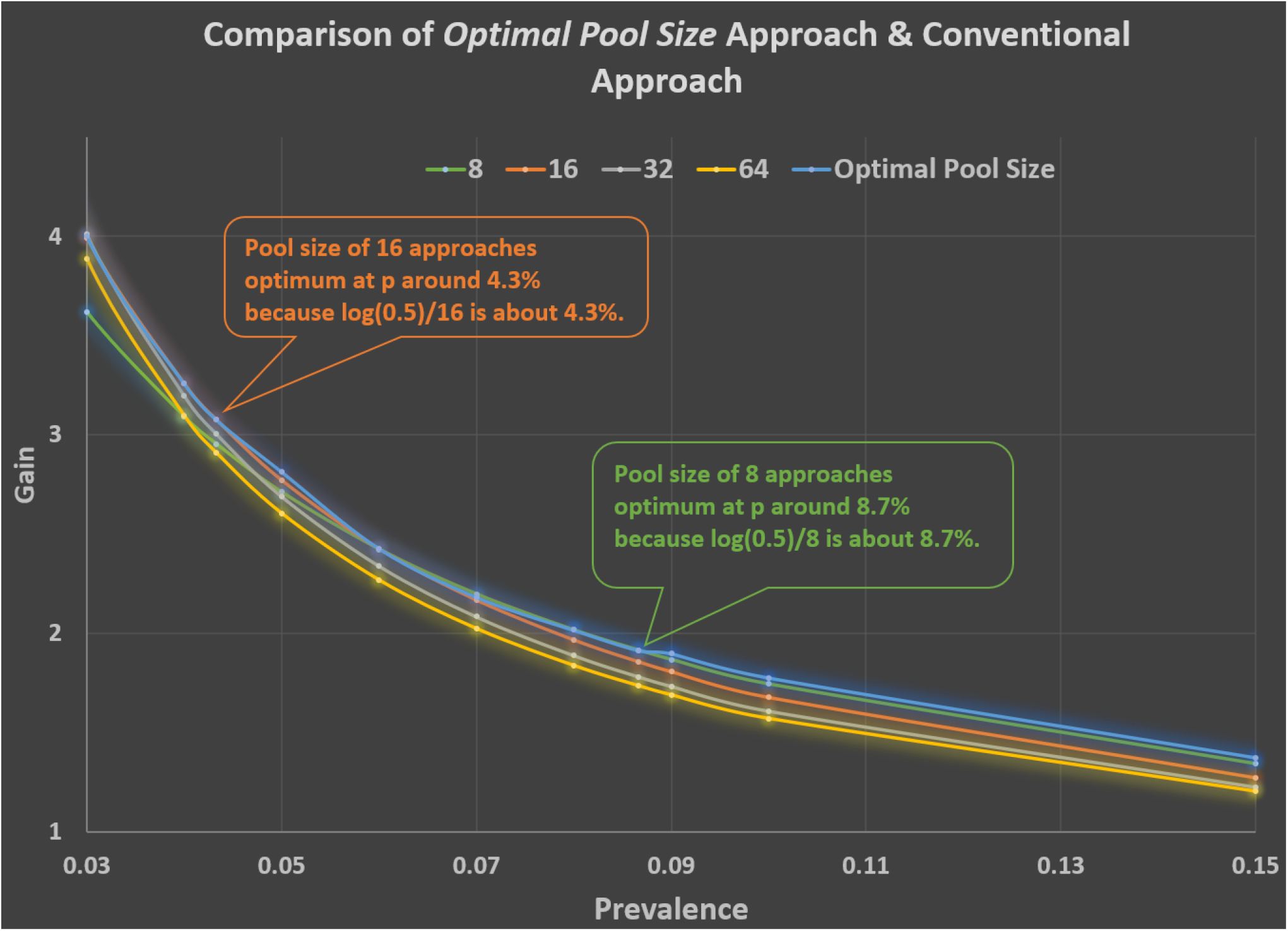
The signatures of the super-even numbers16 and 8 approaching the blue optimal curve can be identified in this zoomed-in version of Figure 6. The blue curve is the Optimal Pool Size model using Eq (9) or Eq (10). A pool size of 8 becomes optimal at *p* around 8% because *log*(0.5)/8 is about 8%. A pool size of 16 becomes optimal at *p* around 4% because *log*(0.5)/16 is about 4%.

## 5. Shifting of Best Compaction (*ϵ*) in High Prevalence (*p*_*i*_) Regime

The ceiling function in the gain formula introduces important modifications. Figure 8 is a graph of Equation 16. It shows the gain dependence on compaction for these scenarios. The cliff-like drops on the peak gain can be as large as 11% when *p* = 2%. The *ϵ* also appears to shift towards 0.4 from 0.5 for larger values of prevalence. These modifications need to be addressed in actual implementation of the optimum pool size approach when the prevalence is ≥ 2%. The data presented in this paper has included the necessary discretization modifications.

**Figure 8.**
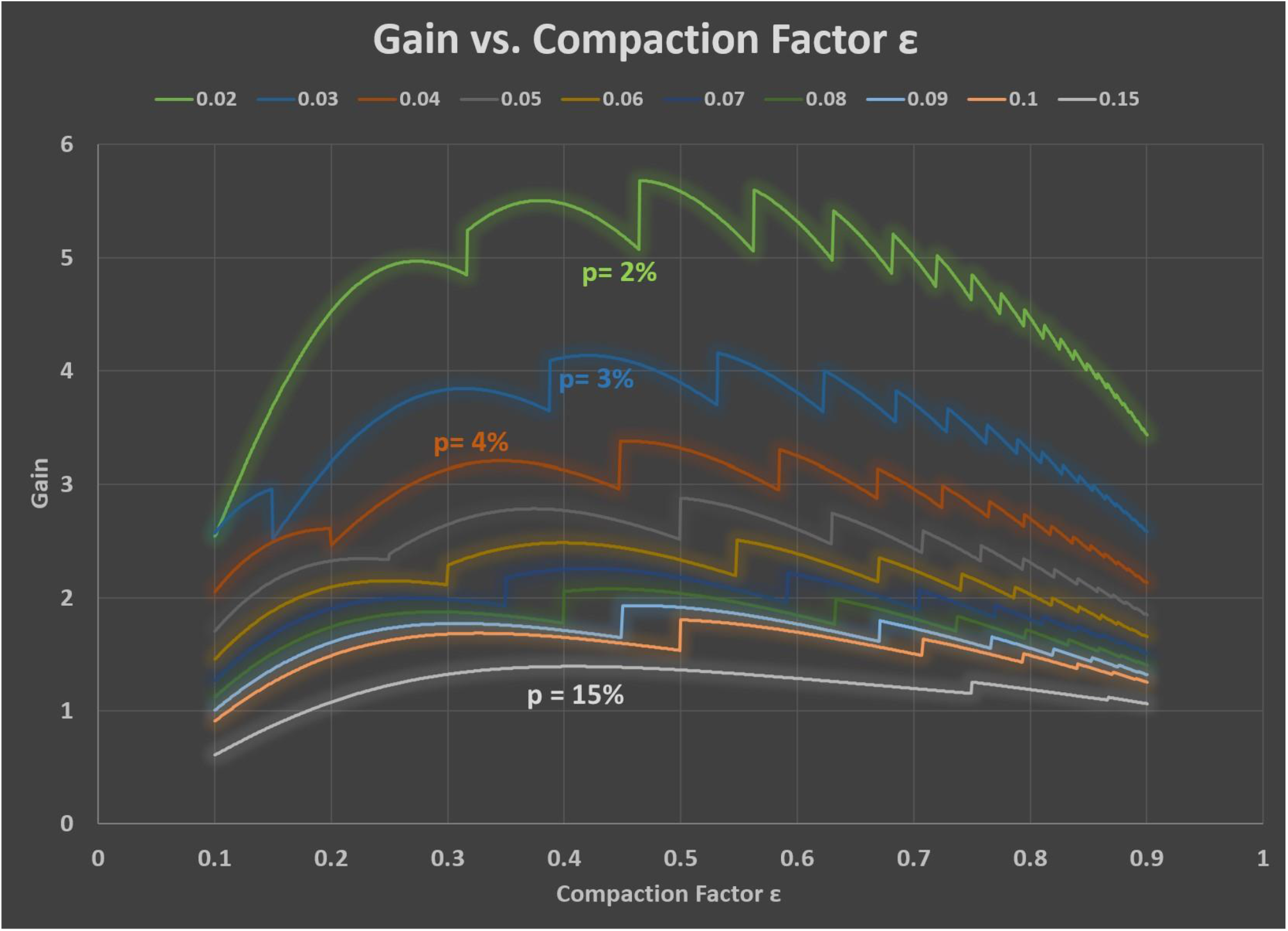
The ceiling function in the gain formula (Equation 1) induces a shift of peak-gain ε from 0.5 to 0.4 when prevalence is higher than 2%. Application of the optimal pool size approach has taken this effect into consideration.

## 6. Discussions and Policy Implications

The potential high gain region of operation can reduce overall testing costs and time. In order to take full advantage of low prevalence, it is essential to conduct smart screening to accomplish a hybrid group testing by forming heterogeneous subgroups. Smart grouping calls for risk assessments. The risks can be measured in a number of ways traditionally. Some recent techniques involve using artificial intelligence, in which a training data set of individual diagnoses and corresponding risk factors are used to establish a binary regression model. This model can then be applied to the current individuals being screened in order to estimate their risk of having the disease.

As an example (See Table 6), when the whole target population is divided into low risk (p = 0.1%), medium risk (p = 1%), and high risk (p = 20%) groups, a gain of 64.7 can be expected for the low risk group and a gain of 9.6 can be expected for the medium risk group. A composite gain of 30 is accomplished in this example.

**Table 6.** An example illustrating a composite gain of 30. With a population of 1,000,000, the number of test required is 33,578.

|  | Group % | Prevalence | # of people | Tests Required |
| --- | --- | --- | --- | --- |
| **Low Risk** | 90% | 0.10% | 900,000 | 14,055 |
| **Medium Risk** | 9% | 1% | 90,000 | 9,523 |
| **High Risk** | 1% | 20% | 10,000 | 10,000 |
|  |  |  | **1,000,000**<br>**People** | **33,578**<br>**Tests Required** |

This idealized mathematical study focuses narrowly on improving the group testing efficiencies. Other important and compelling issues related to testing—such as sensitivity, specificity and technical logistics, have not been discussed here. They are very relevant and deserve to be studied in detail. It is the hope of the authors that the unconventional choices of pool size in this approach will trigger development of future robotic pipetting to accommodate cross-pool mixing arrangement during retest stages. These advanced robots also have to handle rows and columns of non-super-even numbers as well as super-accurate low-dose dispensing.

The approach studied here, deep down, is isomorphic to the conventional multi-stage “halving” algorithms based on super even numbers. However, it offers advantages when the prevalence is initially unknown, or when the prevalence is changing with time. It is believed that the initial choice of pool size is very relevant in retaining a high gain in testing efficiency. Using the usual 16 (or 64) to start group testing, though pleasingly simple, may cost us in efficiency.

## Data Availability

All data is available upon request.

## Footnotes

1* Prevalence is the percentage of persons in a population who have a particular disease (or attribute) at a specified point in time (or over a specified period of time).

